# Meningitis vaccination campaign in the context of COVID-19 in Cameroon

**DOI:** 10.64898/2026.06.02.26354702

**Authors:** Marie Angèle Mbang, Adidja Amani, Fabrice Zobel Lekeumo Cheuyem, Rick Tchamani, Collins Asaah Tatang, Aimé Gilbert Mbonda Noula, Christian Nöel Bayiha, Jeudi Debnet, Zacheus Nanje Ebongo, André Arsène Bita Fouda

## Abstract

**Objective:** The study aimed to describe the challenges, best practices, and lessons learned during meningitis vaccination campaigns conducted in the context of COVID-19 in Cameroon in 2020.

**Results:** During the prevention campaigns, 3,460 individuals were selected. All were tested before the campaign (100%). Eight cases were positive, representing a positivity rate of 0.23% (8/3,460). The campaign was carried out using a fixed strategy in health facilities and prisons and a fixed-temporary strategy in communities. Most health areas received sufficient quantities of COVID-19 equipment for some items and insufficient quantities for others. No screening was done during or after the campaign. The main difficulties encountered were compliance with social distancing and the continuous wearing of gowns. The challenges faced were the screening of actors and the use of personal protective equipment. Lessons learned: aspects related to COVID-19 impacted the speed of the campaign. Vaccination coverage ranged from 91% to 140% in prisons on the one hand, and from 35% to 112% in the health areas surrounding prisons on the other. The campaign in the context of COVID-19 was effective. Compliance with barrier measures was not optimal due to difficulties encountered with aspects such as social distancing, continuous wearing of gowns, screening of participants during and after the campaign, and insufficient personal protective equipment.

## Introduction

Before 2010, approximately 90% of meningitis epidemics were caused by *Neisseria meningitidis* serogroup A (NmA) across the 26 countries of the African meningitis belt, including Cameroon [1–3]. Classic epidemics due to Meningitis A typically occurred every 8 to 10 years. These epidemics generally begin in the middle of the dry and cold season (December–February) and stop spontaneously after 3 to 5 months, at the start of the rainy season (May–June) [4, 5]. In 2010, the city of Ngaoundéré experienced its first epidemic of meningococcal meningitis. The two northernmost regions of Cameroon, the North and the Far North, are considered part of the African meningitis belt and are periodically affected by meningococcal meningitis epidemics [6].

To address this disease, the Heads of State of the African Union signed the Yaoundé Declaration in 2008 to eliminate Meningitis A epidemics [7]. This elimination was to be achieved through three main phases: the organization of initial preventative campaigns, followed by follow-up campaigns to catch up cohorts born after the first campaign, and finally, the introduction of the Men A vaccine into routine immunization [8, 9] .

In Cameroon, as part of this process, 6,725,245 people aged 1–29 years were vaccinated with MenAfriVac between 2011 and 2013 during preventative campaigns conducted in five regions of the country traversed by the meningitis belt: the Adamawa, Far North, North, Northwest, and East regions [2, 10]. Since these campaigns, the distribution of germs causing meningitis in the country has changed, with a 39% decrease in the prevalence of meningococcus A and a shift in the bacteriological profile of meningitis [11]. This change is marked by a predominance of *Streptococcus pneumoniae, Neisseria meningitidis* serogroup X, *Neisseria meningitidis* serogroup C, *Neisseria meningitidis* serogroup W, and *Haemophilus influenzae* type b [12, 13].

Vaccination is an essential element in managing the disease [2]. The World Health Organization (WHO) indicates that approximately 20% of people who contract bacterial meningitis suffer long-term consequences, ranging from disability to sometimes lifelong neurological impairment. These sequelae include hearing loss, which can be particularly detrimental for children during school integration [13]. In addition, approximately 50% of patients die without treatment [14]. Even if the disease is diagnosed early and appropriate treatment is initiated, 5% to 10% of patients typically die within 24 to 48 hours of symptom onset. Meningitis can also cause brain damage or learning disabilities in 10% to 20% of surviving patients [15, 16]. Vaccination is thereby essential and cost-effective tool to prevent deaths from this life-threatening disease [2]. The health authorities thereby prepared and conducted regions-specific campaigns against Meningitis in Cameroon.

The objective of the preventative meningitis vaccination campaign was to strengthen the immunity of the population in and around correctional facilities against meningitis in the context of COVID-19. Public health and social measures (PHSMs) were being implemented worldwide to limit transmission and reduce COVID-19-related mortality and morbidity. These included individual and societal non-pharmaceutical interventions against COVID-19 [17]. PHSMs encompass personal protection measures (such as hand hygiene, respiratory hygiene, mask-wearing); environmental measures (such as cleaning, disinfection, ventilation); and physical distancing measures (such as limiting the size of gatherings and maintaining distance in public or at work) [18]. In general, the common and essential measures for all professional activities include ventilation of workplaces, mask-wearing, distancing, and hand hygiene [19].

The SARS-CoV-2 (COVID-19) pandemic negatively impacted the delivery of health services [20, 21]. The virus first appeared in Cameroon on March 6, 2020 and rapidly progressed to affect the entire country [22]. Cases have been reported in all ten regions of the country though the majority remain in Central and Littoral regions [23]. Given the severity, gravity, and scale of the pandemic, it was necessary to protect all personnel during the campaign. The use of personal protective equipment (PPE) was essential for every person involved in the campaign, particularly in a context of community transmission. The plan for implementing barrier measures had to ensure that exposure risks to the virus were avoided, non-avoidable risks were evaluated, and necessary collective and individual protection measures were identified [17].

The objective of this study was to determine vaccination performance and highlighted the best practices and lessons learned during the first meningococcal meningitis vaccination campaign in the context of COVID-19 in Cameroon.

## Material and methods

### Design and period

This descriptive cross-sectional study examined the mass preventive meningitis vaccination campaign, which was conducted in two rounds from July 26th to September 16th, 2020.

### Sites

The preventive vaccination campaign’s initial phase targeted prison environments and refugee camps in the Far North and North regions between July 20th and 31st, 2020, followed by the central prison of Ngaoundéré in the Adamawa region from August 18th to 30th, 2020. Specifically, the first part of the campaign in the North Region encompassed the prisons of Garoua, Guider, Poli, Tcholiré_1, and Tcholiré_2. In the Far North Region, the campaign focused on the Minawao refugee camp, the prisons of Maroua, Kaelé, Kousseri, Mokolo, Mora, Yagoua, Moulvoudaye, Makary, and Doukoula, and the health areas of Gadala, Zamay, and Gawar, as well as the central prison of Ngaoundéré in the Adamawa Region. The second phase of the campaign took place from September 4th to 16th, 2020, in eight Health Districts (HDs) of the Far North Region: Kaelé, Kousseri, Makary, Maroua 2, Mokolo, Mora, Moulvoudaye, and Yagoua.

### Participants

Eligible participants for the campaign included individuals aged 2 years and older, specifically targeting residents of the identified prison areas and refugee camps, as well as populations within eight Health Districts in the Far North Region. Individuals who were not residents of the prison areas or their immediate vicinity, and those exhibiting signs of prostration and hyperthermia, were excluded from participation.

### Media briefing on the campaign

Given the ongoing COVID-19 pandemic and the proliferation of false information and rumors on social networks that often threaten public health initiatives, especially mass actions, media officials played a crucial role in achieving the objectives of the Meningitis prevention program. To this end, several journalists from television, radio, and the written press participated in a briefing session. The purpose of this briefing was to equip these journalists with accurate information to effectively combat the Meningitis epidemic, particularly within the challenging context of the COVID-19 pandemic.

### Material

Vaccine transport involved moving supplies from the Centre region to all other regions, and subsequently to the designated Health Districts (HDs), utilizing appropriate vehicles and materials specifically designed to maintain the cold chain throughout the entire transportation process. At the operational level, meticulous arrangements ensured the vaccines were stored within the recommended temperature range of +2°C to +8°C until administration, adhering to manufacturers’ guidelines. The HDs managed their vaccine stocks through a rotational replenishment system, with quantities allocated based on the targeted population size and anticipated wastage.

### Data collection method

Following training sessions for HDs management teams, held at the regional delegations of public health, each HD team then trained its own social mobilizers and vaccinators. To adhere to the one-meter social distancing guidelines necessitated by the COVID-19 health context, the training was conducted in smaller groups, supported by central supervisors and partners from the WHO. The training curriculum covered both the theoretical and practical aspects of meningitis, including its definition, clinical manifestations, preventive measures, and the rationale behind the campaign’s preparation. In total, seven hundred and ninety-six vaccinators and social mobilizers were trained prior to the campaign’s launch. Data collection during the campaign utilized tally sheets, and summary sheets, which were employed by the vaccination teams at each participating site and data mask utilized by data manager at the central level.

### Statistical analysis

The data were entered and analyzed using Excel 2026 software. Vaccine coverage was defined as the number of people vaccinated divided by the target population concerned. In the prisons and the concerned refugee camp, the target populations were estimations provided by the officials in charge of these institutions, while in the HDs, the target populations were the 2020 estimates provided by the Ministry of Public Health.

## Results

### Social mobilization

Before the three days of vaccination, social mobilization activities were carried out. For the vaccination campaign’s community mobilization efforts, the objective of informing 100% of household heads in health districts bordering prisons and 100% of officials in charge within the prisons was successfully achieved prior to the arrival of vaccination teams. Population was informed in their household by social mobilization workers. Furthermore, all identified instances of refusal during the campaign were effectively managed.

### Leadership, planning, and communication in the COVID-19 context

During the training of personnel, awareness was raised regarding adherence to barrier measures (respecting physical distancing, wearing masks, using alcohol-based hand sanitizer, and wearing gowns). Before the personnel training, a nasal COVID-19 screening was performed on all participants by laboratory staff working in the health facilities hosting the campaign’s fixed posts. 8 out of 3,460 personnel (0.23%) had confirmed positive rapid diagnostic test results. Those who tested negative were retained, while those who tested positive were replaced and managed according to the national COVID-19 protocol.

### Vaccination coverage

In the Northern region, a vaccination campaign across five prisons (Garoua, Guider, Poli, Tcholliré_1, and Tcholliré_2) showed an overall coverage ranging from 105% to 107%. The 11-18 years age group had the highest coverage at 107%, while the 2-10 and 19+ year groups maintained a 105% coverage (Table 1).

**Table 1.** Vaccination coverage by age groups during the meningitis campaign in the North Region prisons, July 2020 (phase one campaign)

| Site | Target population<sup>1</sup> |  |  |  | Vaccination coverage<sup>1</sup> |  |  |  |
| --- | --- | --- | --- | --- | --- | --- | --- | --- |
|  | n |  |  |  | n (%) |  |  |  |
|  | 2-10 | 11-18 | ≥19 | Total | 2-10 | 11-18 | ≥19 | Total |
| Garoua | 208 | 381 | 1 682 | 2 271 | 237 (114) | 431 (113) | 1832 (109) | 2500 (110) |
| Guider | 45 | 187 | 418 | 650 | 45 (100) | 187 (100) | 418 (100) | 650 (100) |
| Poli | 71 | 42 | 286 | 399 | 71 (100) | 42 (100) | 286 (100) | 399 (100) |
| Tchollire_1 | 114 | 78 | 416 | 608 | 114 (100) | 78 (100) | 413 (99) | 605 (99) |
| Tchollire_2 | 95 | 38 | 187 | 320 | 95 (100) | 38 (100) | 187 (100) | 320 (100) |
| **Total** | **533** | **726** | **2 989** | **4 248** | **562 (105)** | **776 (107)** | **3 136 (105)** | **4474 (105)** |

<sup>1</sup>By age groups in years

Overall vaccination coverage was 71% in the Minawao refugee camp. In contrast, prison vaccination rates were generally higher, exceeding 100% in seven prisons (Maroua, Kousseri, Mora, Yagoua, Moulvoudaye, Makary, and Doukoula), with Kaelé prison at 97% and Mokolo at 91%. Health area coverage was consistently above or equal to 90%, specifically: Gadala (95%), Zamay (90%), and Gawar (98%). The 11-18 years age group showed the highest vaccination rates in both the refugee camp (93%) and the health areas (over 100%) (Table 2).

**Table 2.** Vaccination coverage by age groups and implementation sites during the meningitis campaign in the Far North Region, July 2020 (phase one campaign)

| Site | Target population<sup>1</sup> |  |  |  | Vaccination coverage<sup>1</sup> |  |  |  |
| --- | --- | --- | --- | --- | --- | --- | --- | --- |
|  | n |  |  |  | n (%) |  |  |  |
|  | 2-10 | 11-18 | ≥19 | Total | 2-10 | 11-18 | ≥19 | Total |
| **Refugies Camp** |  |  |  |  |  |  |  |  |
| Minawao | 22 518 | 10 557 | 24 538 | 57 613 | 15311 (68) | 9867 (93) | 15737 (64) | 40915 (71) |
| **Prison** |  |  |  |  |  |  |  |  |
| Maroua | 0 | 0 | 1 528 | 1 528 | 0 | 0 | 1715 (112) | 1997 (130) |
| Kaele | 0 | 0 | 205 | 205 | 0 | 0 | 158 (77) | 200 (97) |
| Kousseri | 0 | 0 | 328 | 328 | 0 | 0 | 290 (88) | 349 (106) |
| Mokolo | 0 | 0 | 417 | 417 | 0 | 0 | 336 (80) | 380 (91) |
| Mora | 0 | 0 | 228 | 228 | 0 | 0 | 174 (76) | 250 (109) |
| Yagoua | 0 | 0 | 501 | 501 | 0 | 0 | 449 (89) | 530 (105) |
| Moulvoudaye | 0 | 0 | 57 | 57 | 0 | 0 | 70 (122) | 70 (122) |
| Makary | 0 | 0 | 50 | 50 | 0 | 0 | 45 (90) | 70 (140) |
| Doukoula | 0 | 0 | 35 | 35 | 0 | 0 | 21 (60) | 50 (142) |
| **Health area/IHC** |  |  |  |  |  |  |  |  |
| Gadala | 2 562 | 1 201 | 2 792 | 6 555 | 2756 (107) | 1283 (106) | 2231 (79) | 6270 (95) |
| Zamay | 4 830 | 2 264 | 5 263 | 12 357 | 4547 (94) | 2342 (103) | 4237 (80) | 11126 (90) |
| Gawar | 6 389 | 2 995 | 6 963 | 16 347 | 6661 (104) | 3747 (125) | 5666 (81) | 16074 (98) |

<sup>1</sup> Age group in year; IHC: Integrated Health Centre

During this second phase of the campaign, vaccination took place in 8 health areas surrounding the prisons: Kaélé, Kousseri, Makary, Maroua 2, Mokolo, Mora, Mouvouldaye, and Yagoua. The 2-10 years age group had the highest vaccination rates across the different health areas, with 6 out of 8 (75%) achieving a vaccination coverage exceeding 100% (Table 3).

**Table 3.** Vaccination coverage by age groups during the meningitis campaign among community of the Far North Region, September 2020 (phase 2 campaign)

| Site | Target population<sup>1</sup> |  |  |  | Vaccination coverage<sup>1</sup> |  |  |  |
| --- | --- | --- | --- | --- | --- | --- | --- | --- |
|  | *n* |  |  |  | *n (%)* |  |  |  |
|  | 2-10 | 11-18 | ≥19 | Total | 2-10 | 11-18 | ≥19 | Total |
| Kaele | 6 065 | 6 355 | 11 629 | 24 049 | 7998 (131) | 6649 (104) | 12460 (107) | 27107 (112) |
| Kousseri | 12 112 | 12 692 | 23 225 | 48 029 | 18763 (154) | 15275 (120) | 17807 (76) | 51845 (107) |
| Makary | 7 861 | 8 237 | 15 073 | 31 170 | 13824 (175) | 7047 (85) | 9930 (65) | 30801 (98) |
| Maroua 2 | 15 250 | 15 979 | 28 517 | 59 746 | 7218 (47) | 5142 (32) | 9094 (31) | 21454 (35) |
| Mokolo | 10 648 | 11 158 | 22 039 | 43 845 | 12176 (114) | 7635 (68) | 9946 (45) | 29757 (67) |
| Mora | 15 650 | 16 399 | 30 008 | 62 057 | 25626 (163) | 17115 (104) | 19937 (66) | 62678 (101) |
| Moulvoudaye | 8 705 | 9 121 | 16 290 | 34 116 | 6842 (78,6) | 4879 (53) | 6042 (37) | 17763 (52) |
| Yagoua | 21 041 | 22 047 | 40 343 | 83 431 | 28297 (134) | 24552 (111) | 29192 (72) | 82041 (98) |

<sup>1</sup> Age group in year; IHC

For all age groups combined, 100% of the target population was vaccinated in the Ngaoundéré urban central prison. The 11–18 years old constituted a significant group (Table 5).

**Table 5.** Vaccination coverage by age groups during the meningitis campaign in the Adamawa Region, August 2020 (phase 1 campaign)

| Site | Target population<sup>1</sup> |  |  |  | Vaccination coverage<sup>1</sup> |  |  |  |
| --- | --- | --- | --- | --- | --- | --- | --- | --- |
|  | *n* |  |  |  | *n (%)* |  |  |  |
|  | 2-10 | 11-18 | ≥19 | Total | 2-10 | 11-18 | ≥19 | Total |
| Prison |  |  |  |  |  |  |  |  |
| Ngaoundéré | 0 | 59 | 1 348 | 1407 | 0 | 59 (100) | 1348 (100) | 1407 (100) |

<sup>1</sup> Age group in year;

## Discussion

Mass vaccination campaigns aimed at preventing or responding to outbreaks of vaccine-preventable diseases and high-impact diseases (VPD/HID) are effective strategies for reducing mortality and controlling diseases. However, many countries had to postpone these vaccination campaigns due to the physical distancing measures implemented to curb COVID-19 transmission [24].

This article describes the vaccination coverage, best practices, and lessons learned during the preventative meningococcal meningitis ACYW vaccination campaign conducted in refugee camps, prisons, and the surrounding health areas in the Adamawa, Far North, and North regions of Cameroon in September 2020. The vaccine used was Men ACWY-PS.

The success of this preventative campaign was recorded in the prisons and refugee camps. In the health areas surrounding the prisons, the administrative coverages varied and are similar to those obtained in the countries of the meningitis belt in 2018 [25]. The campaign targeted people aged 2 years and older.

The campaign was carried out in two phases. The first phase took place in prisons, where the vaccination coverage was 100%, which can be explained by the fact that the prison environment is a closed setting. The second phase was carried out in 8 health areas surrounding the prisons, where vaccination coverages varied from one zone to another. The target population was those aged 2 years and older, and the 2-10 age group registered vaccination coverages over 100% (131%), similar to that recorded during the reactive vaccination campaign conducted in Banalia, Democratic Republic of Congo, in October 2021 (104.6%) [26].

For communication during this campaign, the media were briefed on the COVID-19 pandemic, meningococcal meningitis vaccination, and the proliferation of false information and rumors via social networks. Community mobilization was also carried out, reaching 100% of the households in the concerned health areas and the officials in charge of the prisons. These strategies are similar to those recommended by Sarah Cruickshank and Samantha Grills in the document entitled: *Immunization strategies: Eradicating Meningitis in SubSaharan Africa* [27].

A nasopharyngeal COVID-19 screening was performed on all personnel. Personal Protective Equipment (PPE) was also used. 8 out of 3,460 personnel (0.23%) had confirmed positive Rapid Diagnostic Test (RDT) results. Positive cases were managed and replaced. New actors; tested negative for Covid-19 immediately replaced those who were tested positive. It is similar to what Amani and al. did at 2022 in oral cholera mass vaccination campaign in urban Cameroon during the COVID-19 era [28].

Coordination, planning, social mobilization, and strategies were the main elements implemented for best practices during these campaigns as recommended by the World Health Organization in 2020 [24].

### Challenges, best practices & lessons learned

#### Challenges

This vaccination occurred in the context of the COVID-19 pandemic. No reluctance was recorded in the prisons and refugee camps. In contrast, cases of reluctance encountered in the community were managed by the personnel. The implementation of COVID-19 guidelines slowed the speed of the campaign and increased the time required for implementation during the second phase by three days.

#### Best practices and lessons learned

(1) Social mobilization: The efforts undertaken by social mobilizers and awareness-raising among the prison officials also allowed 100% of households in the zones surrounding the prisons to be reached. The identification of refusal areas made it possible to manage reluctance without ambiguity. (2) Vaccination technique: Given the context in which the campaign was conducted, the screening of personnel before the campaign allowed for the identification of COVID-19-negative personnel, the isolation of positive personnel, and their referral to a treatment center for management. Personal Protective Equipment (PPE) was used by all personnel during the campaign. (3) To reach the targets, the fixed-temporary strategy implemented in the community and prison settings was the most frequently used compared to the fixed strategy implemented in the health facilities.

## Limitations

During this campaign, the personnel were not tested during or after the activity. The personal protective equipment was insufficient in quantity to cover the entire duration of the campaign. Respecting physical distancing, continuous mask-wearing, and wearing the gown were the major difficulties encountered. The data for this campaign were collected by trained community health workers and not by an independent monitoring team to ensure the validity of all data.

## Conclusions

The preventive vaccination campaign against meningitis was a success, with vaccination coverage reaching 100% in prisons and certain health districts surrounding prisons. The COVID-19 pandemic slowed down the campaign. Infection control and prevention were not optimal due to insufficient personal protective equipment (PPE). In such contexts, it would be advisable to produce sufficient quantities of PPE. This would enable stakeholders to address the major difficulties encountered.

## Acknowledgements

This work is the result of a collaborative effort by numerous stakeholders. The authors acknowledge the contributions of the Ministry of Public Health of Cameroon for administrative support. We also address our gratitude to the members and community leaders of the Adamawa, Far North, and North regions of Cameroon for their relentless dedication to this campaign.

## Author contributions

Conception and design: MAM, Administrative support: ZNE and JD. Data collection and analysis: MAM and FZLC; Writing the first draft of the manuscript: MAM, FZLC, and RT; Editing successive drafts of the manuscript: MAM, FZLC; RT, JD, ZNE, and AABF; Final approval of manuscript: MAM, FZLC, RT, JD, ZNE, and AABF.

## Data availability

The dataset used in the present work is available from the corresponding author upon reasonable request.

## Declarations

### Ethical approval and consent to participate

The ethical approval and consent to participate was waived by the IRB authorities of the Ministry of Health as this data study derived from a national campaign.

### Consent for publication

Not applicable.

